# Emergence of Resistance to Fluoroquinolones and Third-Generation Cephalosporins in *Salmonella* Typhi in Lahore, Pakistan

**DOI:** 10.1101/2020.02.12.20020578

**Authors:** Farhan Rasheed, Muhammad Saeed, Nabil-Fareed Alikhan, David Baker, Mohsin Khurshid, Emma V. Ainsworth, A. Keith Turner, Ambereen Anwar Imran, Muhammad Hidayat Rasool, Muhammad Saqalein, Muhammad Atif Nisar, Muhammad Fayyaz Ur Rehman, John Wain, Muhammad Yasir, Gemma C. Langridge, Aamer Ikram

**Affiliations:** Allama Iqbal Medical College & Jinnah Hospital (AIMC&JHL) Lahore, Pakistan; Department of Microbiology, Government College University, Faisalabad, Pakistan; Microbes in the Food Chain, Quadram Institute Bioscience, Norwich Research Park, Norwich, NR4 7UQ UK; University of Sargodha, Sargodha, Pakistan; University of East Anglia, Norwich Research Park, Norwich, NR4 7TJ; National Institute of Health, Islamabad Pakistan

**Keywords:** *Salmonella* Typhi, typhoid fever, XDR, cephalosporin resistance, Lahore, Pakistan

## Abstract

**Background:** Extensively drug-resistant (XDR) *Salmonella* Typhi has been reported in Sindh province of Pakistan since 2016. The potential for further spread is of serious concern as remaining treatment options are severely limited. We report the phenotypic and genotypic characterisation of 27 XDR *S*. Typhi isolated from patients attending Jinnah Hospital, Lahore, Pakistan.

**Methods:** Isolates were identified by biochemical profiling; antimicrobial susceptibility was determined by a modified Kirby-Bauer method. These findings were confirmed using Illumina whole genome nucleotide sequence data. All sequences were compared to the outbreak strain from southern Pakistan and typed using the *S*. Typhi genotyping scheme.

**Findings:** Twenty-seven XDR *S*. Typhi isolates were identified from patients at Jinnah Hospital in Lahore between January and April 2019. All isolates were confirmed by sequence analysis to harbour an IncY plasmid and the CTX-M-15 ceftriaxone resistance determinant. All isolates were of the same genotypic background as the outbreak strain from Sindh province.

**Summary:** We report the first emergence of XDR *S*. Typhi in Punjab province of Pakistan confirmed by whole genome sequencing.

**Funding:** This work was supported by the BBSRC-funded QIB Institute Strategic Programme “Microbes in the Food Chain” BB/R012504/1 and its constituent projects Microbial Survival in the Food Chain (BBS/E/F/000PR10349) and Research Infrastructure (BBS/E/F/000PR10352), and the BBSRC funded Core Capability Grant (project number BB/CCG1860/1).

**Research in context:** *Evidence before this study:* Typhoid fever is endemic in Pakistan, with widespread resistance to first line drugs: ampicillin, co-trimoxazole, and chloramphenicol, and to fluoroquinolones. In 2017 the first report of additional resistance to ceftriaxone was published as XDR typhoid fever from southern Pakistan. Treatment of XDR typhoid fever has been a clinical challenge; options are scarce, and the level of chronic carriage is unknown. Current treatment is dependent upon azithromycin and this represents a major threat for the community as resistance has been reported from Bangladesh.

*Added value of this study:* This study is the first genomic report of XDR typhoid fever in a central part of Pakistan, describing the same genetic background as strains responsible for XDR typhoid in Sindh province. We have confirmed that the outbreak strain of XDR *S*. Typhi is now present in the most populated province and second largest city of Pakistan.

*Implications of all the available evidence:* The present study highlights the gravity of the situation - the spread of this strain is of serious concern. The clinical situation remains unchanged since this study was performed; we believe it is imperative that future research focuses upon the biology, transmission and control of this strain.

## Background/Introduction

Typhoid fever, caused by *Salmonella* Typhi, is a global health concern. The burden of typhoid has been estimated at 13·5 million cases resulting in 135,000 deaths annually, with a global incidence of 2·14 in 1000 ^1,2^. In Pakistan, the precise burden is not known but active surveillance studies in Karachi report 4·7 cases per 1000 per year. Travel-associated typhoid fever is also of concern in high income countries with more than 150 cases of typhoid fever annually in England and Wales since 2008 (PHE report 2018); most of these cases have a history of travel to south Asia. The treatment of typhoid was changed by the emergence of multi-drug resistance (MDR) in *Salmonella* Typhi, which rendered first line antibiotics (amoxycillin, co-trimoxazole, and chloramphenicol) and fluoroquinolones ineffective ^3^. The treatment of choice became a third-generation cephalosporin, most commonly ceftriaxone ^4^. In 2016, there was a report from Karachi of the emergence of extensively drug resistant (XDR) *S*. Typhi ^5^. The *S*. Typhi strains described as XDR are MDR strains with additional fluoroquinolone and ceftriaxone resistance, leaving azithromycin, piperacillin-tazobactam, or carbapenem as the treatment options ^5^. While a report of fluoroquinolone or ceftriaxone resistance in the Enterobacteriaceae is not uncommon in Pakistan, the isolation of *S*. Typhi harbouring resistance to these antibiotics is much more unusual. The current study was prompted by the blood culture isolation of XDR *S*. Typhi from patients under investigation at Jinnah Hospital, Lahore, raising the possibility that the Karachi outbreak strain was circulating in Lahore. Here we investigated 27 *S*. Typhi isolates obtained in less than 4 months and compared their whole genome sequences with the XDR outbreak strain.

## Methods

This study was carried out at the Department of Pathology, Jinnah Hospital, Lahore, Pakistan and Quadram Institute Bioscience, Norwich, United Kingdom.

### Isolation and identification of isolates

Blood cultures were collected, following normal diagnostic protocols, from patients with fever admitted to the Jinnah Hospital Lahore, Pakistan. A Gram stain was performed directly from blood culture broth; the broth was inoculated on blood agar and MacConkey agar. Bacterial colonies were identified biochemically using API 10 or API 20E (BioMérieux, France). Isolates were confirmed as *Salmonella* Typhi by serological reactions to O, H and Vi antigens (Pro-Lab Diagnostics, Canada). Antibiotic susceptibility testing was performed using a modified Kirby-Bauer method according to Clinical Laboratory Standard Institute (CLSI) 2019 guidelines.

Twenty-seven XDR *S*. Typhi isolates were collected between January 2019 and April 2019 and archived at -80 °C. The isolates were re-cultured on blood agar and DNA was extracted for sequencing from single colonies.

### DNA extraction

DNA extraction of *S*. Typhi isolates was carried out using a modified protocol of the PuriSpin Fire Monkey kit (RevoluGen, UK). In brief, 1 ml overnight *S*. Typhi culture was harvested and cells were resuspended in 100 µL of lysozyme (3 mg/mL), 1·2 % Triton X-100, and incubated at 37 °C, 180 rpm for 10 min. 300 µL of lysis solution and 20 µL of Proteinase K (20 mg/mL, Qiagen, UK) was added to the partly lysed cells and incubated at 56 °C for 20 min. After lysis, 10 µL of RNase A (20 µg/µL, Sigma) was added to the suspension and incubated for a further 10 min at 37 °C. Binding solution 350 µL and isopropanol (75 %) 400 µL were added to the lysed cells and the lysed cells were transferred to the spin column. Bound DNA was washed as per manufacturer’s instructions and eluted in 2×100 µL of elution buffer. DNA was shipped to Quadram Institute Bioscience, Norwich, UK and concentrations were determined using the Qubit dsDNA broad range assay kit (Thermo Fisher, UK) on a Qubit 3·0 Fluorometer (Thermo Fisher).

### Whole genome nucleotide sequencing

Genomic DNA concentrations were adjusted to 0·5 ng/µL with nuclease-free water for sequencing. A tagmentation mix of 3 µL was mixed with 2 µL of genomic DNA (0·5 ng/µL) and incubated at 55 °C for 10 min in a PCR block. Kapa2G PCR master mix was added 11 µL to each well sample in a 96-well plate alongside 2 µL of each of P7 and P5 of Nextera XT Index Kit v2 index primers (Illumina). The PCR was carried out following standard Illumina guidelines. The DNA fragment libraries were quantified using the Quant-iT dsDNA high sensitivity assay kit (Thermo Fisher) and pooled. The final pool was double-SPRI size selected between 0·5 and 0·7X bead volumes using KAPA Pure Beads (Roche) and the pool was run at a final concentration of 1·8 pM on an Illumina Nextseq500 instrument using a Mid Output Flowcell (NSQ® 500 Mid Output KT v2(300 CYS), Illumina). Data was uploaded to Basespace (www.basespace.illumina.com) where the raw data was converted to 8 FASTQ files for each sample.

### Genome assembly and annotation

Genome nucleotide sequences of all strains were determined using 150 bp paired-end sequencing on the Illumina Nextseq500 platform. All raw sequence data have been deposited in the NCBI Sequence Read Archive (SRA) under BioProject accession PRJNA602421. Sequenced reads were screened using Centrifuge v1.0.4 to confirm the species of each sample as *Salmonella enterica* ^6^. *De novo* assembly of individual genomes was carried out using Shovill version 1.0.0 (https://github.com/tseemann/shovill, accessed November 2019), which internally corrected sequencing errors, performed genome assembly using SPAdes ^7^, and removed erroneous contigs. Assembly quality was assessed via QUAST v5.0.2 ^8,9^. Draft genomes were annotated using Prokka v1.14 ^10^.

### Genotyping and antimicrobial gene prediction

*In silico* Multi-Locus Sequence Typing was predicted using “MLST” v2.17.5 (https://github.com/tseemann/mlst, accessed November 2019) using the *Salmonella* MLST database hosted by EnteroBase ^11^. To calculate the genotype using the Genotyphi scheme ^12^, draft genomes were aligned using Parsnp v1.2 ^13^ to the Typhi CT18 reference genome (Accession no. AL513382.1) and resulting variant calls (vcf) were run through Genotyphi (commit version fd1d58b) (Holt, 2019), as described on the GitHub repository. Antimicrobial resistance genes were detected using ARIBA v2.14 ^14^ with default parameters.

### Phylogenetic analysis

Ninety-two additional *S*. Typhi genomes were included to provide context to the 27 from this study ^5^. The outgroup and reference genome (ERL12148, accession no. LT883153.1) was selected using the following criteria; the strain was closely related (i.e. *S*. Typhi haplotype H58) and the strain was not associated with the 2016 outbreak. Single nucleotide polymorphisms (SNPs) were identified against the reference using Snippy v4.2.1 (https://github.com/tseemann/snippy, accessed November 2019) and visually inspected in Artemis ^15^. Gubbins v2.3.1 ^16^ was used to define non-recombinant SNPs, which were used with RaxML ^17^ to construct the final phylogeny. A reference IncY plasmid (Pak60006-2016)^5^ was used with Blast Ring Image Generator (BRIG) ^18^ to identify nucleotide identity between the reference genome and strains genome sequenced as part of this study.

### Role of the funding source

The funders had no role in study design, in the collection, analysis and interpretation of data, in the writing of the report or the decision to submit this paper for publication.

## Results

### Phenotypic findings

Although strict surveillance data is not available for typhoid fever in Lahore, a 12-fold increase in isolation rate was observed at Jinnah Hospital over the 12-month period July 2018 to June 2019 (number of cases = 370) relative to the previous 12 months (n<30). Twenty-seven isolates of *S*. Typhi were isolated from the blood of consecutive patients between 14^th^ January and 1^st^ April 2019. Fourteen isolates were from males and thirteen from females. The age range varied from 15 months to 28 years, with 11 of the isolates from under-fives, 13 from 5-16 years old and 3 from over-sixteens (Supp Table S1). This was a typical sample of patients presenting to Jinnah Hospital with suspected typhoid fever. All of the isolates were phenotypically and biochemically identified as *S*. Typhi and serologically confirmed as positive to O, Vi and H antibodies. Antibiotic susceptibility testing showed that these isolates were susceptible only to azithromycin, carbapenems and piperacillin-tazobactam (Supp Table S2). All were resistant to the standard treatment for typhoid fever including ceftriaxone. For all patients, treatment was started with intravenous meropenem followed by azithromycin. There was no mortality reported due to XDR *S*. Typhi.

### Genotypic analysis

Whole genome nucleotide sequences revealed that all isolates were sequence type (ST)1 and identified as 4.3.1.1.P1 under the Genotyphi scheme, the same as the strains from the 2016 outbreak reported in Sindh province ^5,12^. We compared our recent Lahore isolates to ∼100 *S*. Typhi genomes from southern Pakistan and generated a maximum-likelihood phylogenetic tree from 68 filtered and neutral SNPs (Figure 1). All the XDR isolates differed by 6 SNPs from the MDR *S*. Typhi clade, in accordance with findings from the 2016 outbreak ^5^. Our Lahore isolates also intermingled with those from the 2016 outbreak, indicating few, if any, SNP differences between these strains. It is plausible therefore that all the XDR isolates originated from a single strain that acquired the XDR phenotype. Within the XDR clade, the Lahore isolates formed two clusters on the tree, one consisting of isolates LAH17, 5, 24 and 23, and the other of LAH2, 26, 29, 18, 11, 21, 22, 25, 15, 19, 8, 10, 6, 13, 4 and 20, suggesting patient exposure to point sources of infection.

**Figure 1.**
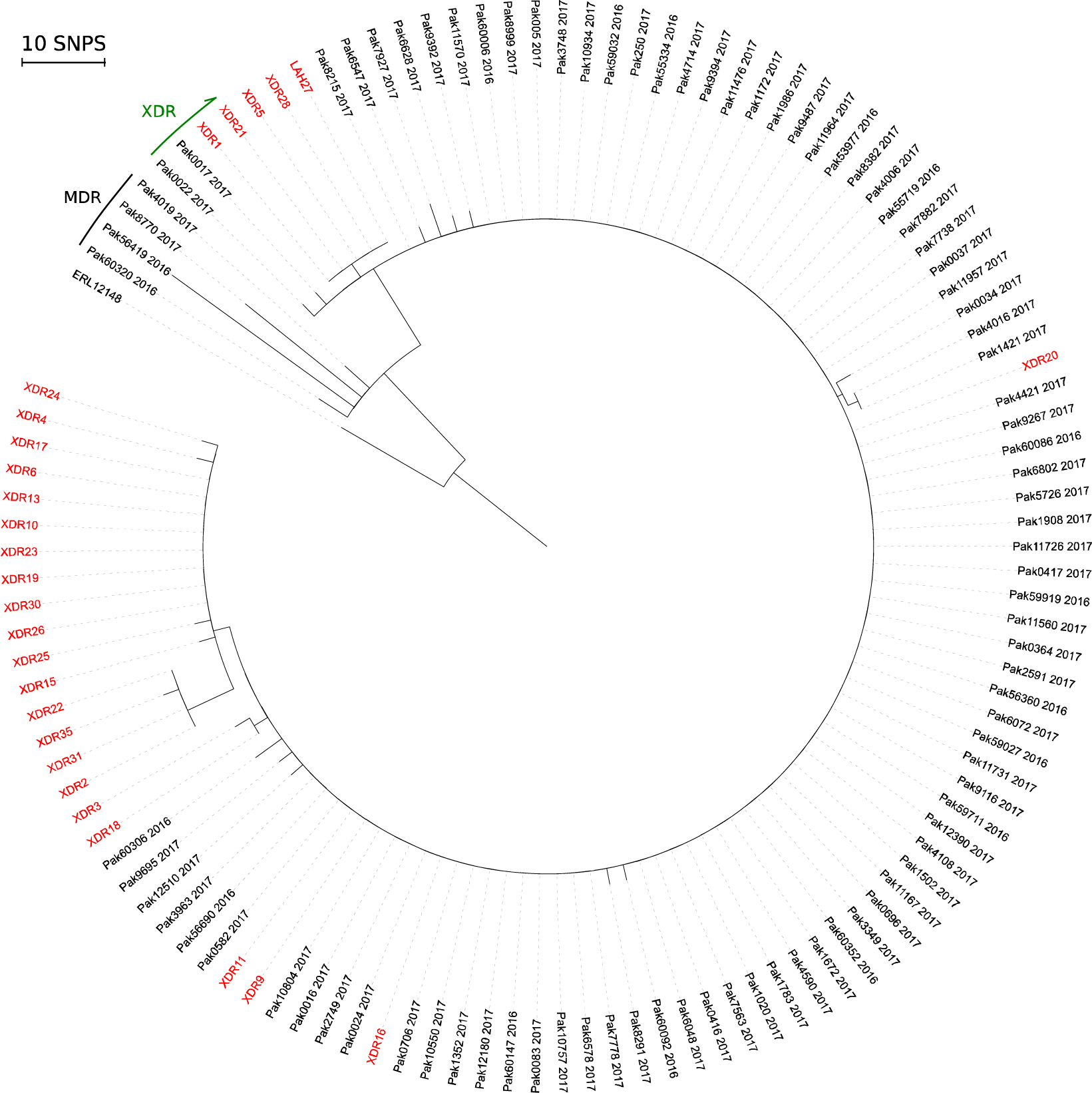
Phylogenetic tree of XDR *S*. **Typhi** Radial phylogram of maximum-likelihood phylogeny (RaxML) rooted to *S*. Typhi ERL12148 (accession LT883153.1). PAK strain numbers in black are from Klemm et al (2019) and LAH strain numbers (red) are from Lahore (this study). Black bar labelled MDR, indicates MDR strains and green arrow indicates that all strains from PAK0002 clockwise to LAH20 are XDR.

Analysis of all 27 draft genome sequences from Lahore indicated the presence of both an IncY plasmid and the CTX-M-15 determinant associated with ceftriaxone resistance. The CTX-M-15 resistance determinant was located on an IncY plasmid in the southern Pakistan outbreak strains; our BRIG analysis suggested that this was also the case for the Lahore isolates (Figure 2).

**Figure 2.**
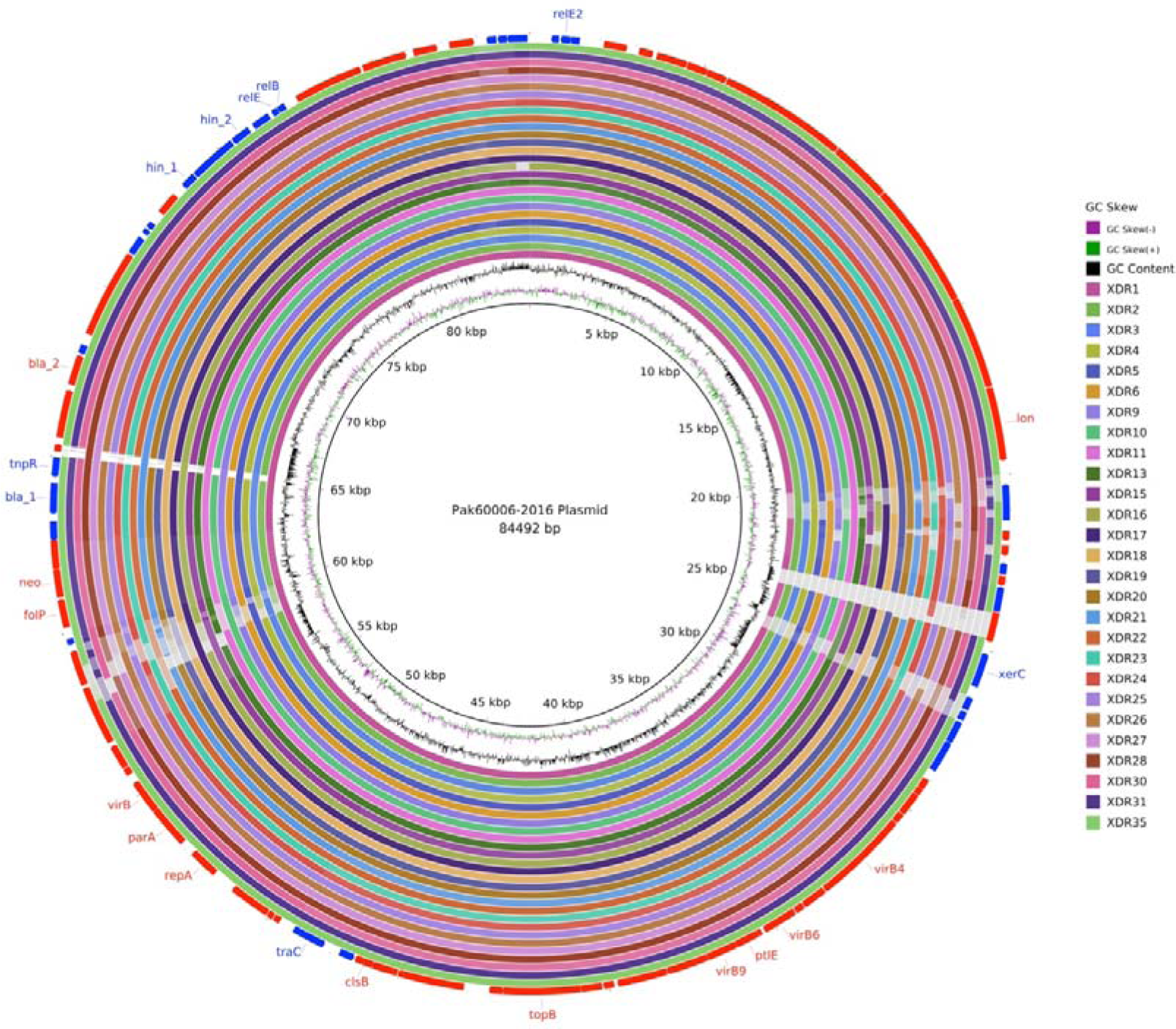
CTX-M-15 located on IncY plasmid. The inner circle represents the reference plasmid sequence p60006. Concentric rings indicate BLAST identity between reference and strains sequenced as part of this study. BLASTn matches between 90 % and 100 % nucleotide identity are coloured from lightest to darkest shade, respectively. Matches with less than 80 % identity, or plasmid regions with no BLAST matches, appear as blank spaces in each ring. Inner most rings plot GC Skew and GC content. Outermost ring shows locations of coding sequences (red - sense; blue - antisense) and gene names are labelled where known. The CTX-M-15 gene is labelled in black.

## Discussion

Here we report *S*. Typhi resistant to amoxycillin, co-trimoxazole, chloramphenicol, ciprofloxacin, and ceftriaxone from Lahore, Pakistan. Our phenotypic and genotypic analysis shows that the *S*. Typhi XDR isolates were indistinguishable from the outbreak strain described in southern Pakistan, indicating that the cases in Lahore are part of the same outbreak. We have investigated a single tertiary health care facility in Lahore but alongside other reports, we believe it is spreading locally ^19^. The isolates from Jinnah Hospital are closely related but several of them harbour SNP differences. Since *S*. Typhi is a clonal pathogen the presence of these SNPs suggest that either the XDR strain is not a recent introduction to the area or that that it has been imported several times ^20^.

The increased level of *S*. Typhi isolation from blood culture and the presence of XDR *S*. Typhi is of serious public health concern. Treatment of XDR *S*. Typhi is one of the growing challenges for infectious disease physicians in Lahore and more widely in Pakistan. The only treatment options that remain in this situation are azithromycin, meropenem, and piperacillin/tazobactam. The treatment strategy for XDR typhoid is intravenous meropenem for seven days followed up by oral azithromycin; this is being used in Pakistan as well as in internationally exported cases ^21^ (Asst. Prof. Fahad Aman Khan, personal communication). Carbapenems are a potent class of antibiotics and an effective treatment for typhoid fever but the presence of carbapenemase-producing Enterobacteriaceae is a critical threat ^21,22^. In addition, the cost of carbapenems in Lahore is prohibitive (one course costs ∼50,000 PKR, equivalent to ∼ 320 USD, January 2020). In practical terms, this leaves many patients with azithromycin as their only viable option (Asst. Prof. Fahad Aman Khan, personal communication).

Furthermore, of the remaining treatment options for XDR *S*. Typhi, only azithromycin is administered orally but the danger is that its use is also prone to the emergence of resistance ^23^. Resistance has already been reported in typhoidal *Salmonella* from Bangladesh, where resistance was conferred by single nucleotide changes in the *acrB* efflux system ^24^ and Pakistan ^25^, and the macrolide resistance genes *mphA, mphB* and *mefB* ^26^ have been reported as conferring resistance in non-typhoidal *Salmonella*. It is likely only a matter of time before one of these mechanisms emerges in the XDR background, rendering carbapenems the only, more expensive, treatment option. It is also worth noting that while azithromycin remains a viable treatment in susceptible XDR isolates, it has been contraindicated for patients on arrhythmic, antipsychotics and citalopram medications leading to an increased risk of cardiovascular-related mortality ^27^.

In conclusion, the risk of single point mutations conferring resistance to azithromycin and of the horizontal transmission of carbapenemase-mediated resistance makes the ongoing threat of XDR typhoid fever very real in Lahore. Epidemiological studies and functional genomics are urgently needed to inform local, national and international control measures and future public health strategies.

## Data Availability

Supplementary Material available
Table S1. Anonymised patient data
Table S2. Antimicrobial susceptibility testing results

## Acknowledgements

The authors gratefully acknowledge the ward and laboratory staff at Jinnah Hospital, Lahore and the support of the Biotechnology and Biological Sciences Research Council (BBSRC). EVA, DB, AKT, JW, MY and GCL were supported by the BBSRC Institute Strategic Programme Microbes in the Food Chain BB/R012504/1 and its constituent projects Microbial Survival in the Food Chain (BBS/E/F/000PR10349) and Research Infrastructure (BBS/E/F/000PR10352). NFA was supported by the BBSRC funded Core Capability Grant (project number BB/CCG1860/1). DNA extraction kits were provided free of charge by RevoluGen.

## Authors’ contributions

FR, MFUR, MY, GCL, AAI, MHR, MSA, MSaq, MAN, MK and AI conceived the study. EVA and DB performed the genome sequencing. NFA, MY, AKT, JW and GCL analysed the data. FR, MFUR, MY, JW and GCL drafted the manuscript. MSae, MAN, MSaq and MK performed the DNA extractions. All authors read and approved the final draft.

## Ethics statement

Ethical approval for this study was obtained from the ethical review board of Allama Iqbal Medical College/Jinnah Hospital, Lahore. All patient data was anonymised to remove any identifying information.

## Declaration of interests

GCL has received fees from RevoluGen for consultancy work.

## Supplementary Material

**Table S1. Anonymised patient data**

**Table S2. Antimicrobial susceptibility testing results**

## Notes

### Competing Interest Statement

The authors have declared no competing interest.

